# Human infection incidence for West Nile virus and Usutu virus in the Netherlands: evidence from a longitudinal nationwide cohort, 2020-2023

**DOI:** 10.64898/2026.10.13.26365405

**Authors:** Marloes Grobben, Coralie Valle, Johan Reimerink, Gert-Jan Godeke, Sandhya Shrestha, Corine Nellestijn, Hanae Abba, Dirk Eggink, Eelco Franz, Bettie C.G. Voordouw, Fiona R.M. van der Klis, Eric R.A. Vos, Gerco den Hartog, Chantal B.E.M. Reusken

## Abstract

**Background:** Ecological surveillance indicates (regional) establishment of West Nile virus (WNV) and Usutu virus (USUV) in the Netherlands. However, the incidence of human infections remains unknown. We investigated the incidence of WNV and USUV infections in the Netherlands in 2020-2023.

**Methods:** We applied a two-tiered serology approach based on protein micro-array and virus neutralization on the participants of the PICO cohort,. Which is a well-described, longitudinal, nationwide population-based cohort. IgG data were used to estimate overall seroprevalence and infection incidence as the primary outcome, and to study antibody longevity. Study set-up enabled tracking of the period of seroconversion and linking of self-reported travel history. The analyses took account of the survey design and weights were incorporated to match the Dutch population distribution following census data.

**Findings:** Among 4281 participants, we identified 113 probable WNV and/or USUV cases, including 55 seroconversions. Thirty-six cases were confirmed by antibody neutralization, of which 12 seroconverted. Six confirmed seroconverters were identified as autochthonous infections. The overall weighted seroprevalence was estimated at 0.54% (0.21-0.87%) for WNV and 0.25% (0.09-0.40%) for USUV. The weighted cumulative incidence in 2020-2023 for autochthonous WNV and USUV infection was estimated as respectively 0.06% (95% CI 0-0.12%) and 0.04% (95% CI 0-0.09%).

**Interpretation:** The estimated infection incidence and seroprevalence of WNV and USUV suggest a substantial underdiagnosis of clinical cases in the Netherlands in 2020-2023. These observations highlight the need for increased awareness among physicians to identify cases and the need for continued paired syndromic-laboratory surveillance for these viruses.

**Funding:** This study was funded by the ministry of Health, Welfare and Sports, the Netherlands.

**Research in context:** *Evidence before this study:* Human West Nile virus (WNV) infections have occurred with increasing frequency and magnitude in various regions of Europe in the past decade, and the virus is still spreading to new areas. Usutu virus (USUV) has been circulating in Europe for decades. To get an overview of the existing evidence on WNV and USUV infections and circulation in the Netherlands, we searched PubMed for studies published in English from database inception until November 17, 2026 using search terms “west nile virus” OR “usutu virus” AND “the Netherlands”. We also reviewed reports from the Dutch National Institute for Public Health and the Environment (RIVM) and the European Centre for Disease Prevention and Control (ECDC). At the start of our study, only eight autochthonous confirmed human WNV infections were detected in the Netherlands, all in 2020. No human cases were reported since while ecological data from wild birds, horses and mosquitoes pointed at local circulation. USUV is known to circulate in the Netherlands since 2016, but evidence of the occurrence of human infections is limited to a single study among blood donors. There were no data on nationwide human infection incidence and seroprevalence in the Netherlands and no data on USUV IgG longevity in humans in general.

*Added value of this study:* This study is the first comprehensive and nationwide longitudinal serological study to investigate both human WNV and USUV infection in the Netherlands. It uses an extensively validated multiplex serological tool in combination with specialized virus neutralization techniques in a cohort representative of the general population. The longitudinal approach enabled tracking of seroconversion in time, and the study design supported the identification of autochthonous cases and linkage to symptom data. The unique study set-up enabled the estimation of the overall seroprevalence for WNV and USUV in the general Dutch population in fall/winter 2023 and the weighted local cumulative infection incidence for the period 2020-2023 to provide insight into the level of underreporting of human cases. Furthermore, the longitudinal follow-up of individual participants provided more insight into the longevity of WNV-specific IgG responses and provided previously unknown information on the longevity of USUV IgG in humans.

*Implications of all the available evidence:* This study is the first to demonstrates the occurrence of unrecognized, autochthonous human infections with WNV and USUV in the Netherlands in the period 2020-2023. While these viruses have a low symptomatic rate in humans, the estimated infection incidence and overall seroprevalence in this study suggest a significant underdiagnosis of (severe) clinical cases, especially for WNV. Our observations will strengthen the awareness among clinicians to include WNV (and USUV) in their differential diagnosis of patients with neurological complaints of unknown aetiology and underline the increasing need to create awareness among the general public for prevention of mosquito bites, which is highly relevant also given the resurgence of clinical cases and fatalities in 2026. This study complements sporadic ecological evidence of local circulation of both WNV and USUV in the Netherlands and highlights the growing relevance of continued WNV and USUV surveillance. The data on the longevity of USUV IgG informs an accurate interpretation of serological diagnostic tests in the context of orthoflavivirus cross-reactivity.

## Introduction

The increasing circulation and expanding geographic distribution of arboviruses such as those in the genus orthoflavivirus, family *Flaviviridae*, cause a growing threat to human health.

West Nile virus (WNV) is an orthoflavivirus and is the most widely distributed arbovirus in humans with endemicity in the Americas, Europe, Africa and Asia.^1^ In Europe until November 2026, human cases have been reported by 23 countries, but some evidence for circulation exists for two additional countries (United Kingdom and Poland) based on molecular data from birds, mosquitoes or equids.^2–4^ In addition, Denmark recently reported serological evidence for local WNV circulation in horses.^5^ Over the past decade, regional outbreaks in Europe occurred with increasing frequency and magnitude. The average yearly number of human cases is around 800, with fluctuation between years and a low point of 164 cases in 2021.^6^ In 2026, reported case numbers exceed all previous years with 2029 human cases in 235 areas across 20 European countries by 11 November 2026.^7^ In the Netherlands, the first human autochthonous infections were observed in 2020, with eight clinical human cases identified.^8,9^ Sporadic observations in resident birds, horses and mosquitoes point at local enzootic circulation of WNV in the following years.^10^ WNV infection is a mandatory notifiable disease in the Netherlands and in 2026, 73 cases have been identified as of 4th November.^4^ While West Nile virus lineage 1 is the most widely distributed, lineage 2 is predominantly found in Europe and is the circulating lineage in the Netherlands.^11^

Usutu virus (USUV) is closely related to WNV and is endemic to Africa and Europe. USUV has been circulating in Europe for at least 3 decades. Over 100 human cases of USUV infection have been identified in Europe, including 30 patients that experienced neurological symptoms.^12^ USUV was detected for the first time in the Netherlands in birds in 2016, when a widespread multi-country avian outbreak occurred in Northwestern Europe,^13^ and can now be considered endemic.^10^ Evidence for the occurrence of human USUV infections comes from a study in blood donations in 2018 and a serological study in bird ringers in 2021.^14,15^

Acute WNV and USUV infections can be detected by RT-PCR, however, the window to apply this method is usually narrow due to the short-lived and low viremia.^16^ Therefore, serology is the most used method for routine diagnostics. WNV-specific IgM typically appears 3-8 days after symptom onset and persists for 1-3 months, while WNV-specific IgG seroconversion occurs about at median eight days after symptom onset and may persist for several years after infection.^17^ However, even compared to other orthoflaviviruses, USUV and WNV share high amino acid sequence similarity in their structural proteins. This antigenic similarity leads to antibody cross-reactivity, which complicates serological diagnosis.^18^ Cross-reactivity between related orthoflaviviruses could originate from previous orthoflavivirus infections or orthoflavivirus vaccination. Therefore, immunoassays are typically confirmed with comparative virus neutralization assays, which is considered the gold standard method when the specific aetiology is important for subsequent actions like public health control measures.^16^

The incidence of WNV and USUV infections is likely underreported across the world due to the high percentage of asymptomatic (∼80%) and mild infections (∼20%), lack of health care provider awareness, and limitations in diagnostic testing. Worldwide, over 50% of persons with viral encephalitis remain undiagnosed,^19^ which is likely similar in the Netherlands. While both USUV and WNV seem (regionally) established in the Netherlands based on animal and mosquito surveillance, and the first human clinical cases since 2020 appeared in 2026, the incidence of human infections is unknown. Here, we used a two-tiered multiplex serology approach to study the seroprevalence and cumulative infection incidence for WNV and USUV, IgG antibody kinetics and the occurrence of local transmission to humans, in a unique, longitudinal, nationwide, population-based cohort in the period 2020-2024 which includes four years with no human cases reported.

## Methods

### Study cohort

The PIENTER Corona (PICO) serosurveillance study is a nationwide longitudinal sample of Dutch citizens across all ages and regions/municipalities initiated by the National Institute for Public Health and the Environment of the Netherlands (RIVM).^20–22^ The PICO-study protocol was approved by the Medical Ethics Committee MEC-U, the Netherlands (Clinical Trial Registration NTR8473), and conformed to the principles embodied in the Declaration of Helsinki. All participants provided written informed consent. Participants self-collected fingerstick blood samples and completed questionnaires over a total of 13 consecutive rounds, starting March 2020 (PICO-1) and ending December 2024 (PICO-13). The time windows of the study rounds are shown in Appendix 1 p 7. Questionnaires addressed, among others, sociodemographic characteristics, existing comorbidities, occurrence of symptoms related to respiratory disease and job category (Appendix 2 and 3). Fingerstick blood samples were collected by participants in tubes, shipped to the RIVM by medical priority mail, where serum was processed and subsequently stored at -20°C. All participants were included based on random-selection of the up-to-date Dutch population registry, and sampled proportional to the inhabitant size of the municipality, using 17 pre-defined age strata. A total of 3233 participants were included in PICO-1.^20^ All these participants had previously participated in the 2016/2017 nationwide PIENTER-3 serosurveillance study and provided consent to be contacted for follow-up research.^23^ The cohort also included a smaller subset of participants specifically from areas with a lower childhood vaccination coverage. The cohort was supplemented twice with additional participants in PICO-2 and PICO-6.^21,22^ Over 10,000 unique participants were included throughout the study, and 4587 participants remained in the last sampling round in 2024 (PICO-13). We started our analysis with screening of PICO-11, the most recently completed round at that time, which took place in fall/winter 2023 (Appendix 1 p 7). In PICO-11, 4977 participants donated a serum sample, the baseline characteristics are shown in Appendix 1 p 2.

### Production and purification of protein antigens

Non-structural protein 1 (NS1) antigens of WNV lineage 2 (GenBank ID AY532665.1 (strain B956) and WZE65508.1 (the Netherlands)), WNV lineage 1 (GenBank ID AHH02658.1 (NY99)) and USUV (GenBank ID AQM55263.1 (Flemalle/2016)) were produced as previously described.^24^ Briefly, expression plasmids (produced by Genscript, Rijswijk, the Netherlands) containing the protein gene followed by a HisTag were transfected in HEK-293F cells maintained in FreeStyle™ 293 Expression Medium (Gibco, Thermo Fisher Scientific, Breda, the Netherlands) using Polyethylenimine Hydrochloride “Max” (PEI MAX) (Polysciences, Bergstrasse, Germany). Supernatant was harvested after 7 days by centrifugation. Secreted proteins were purified from the filtered supernatant by binding to Ni-NTA beads (Chelating Sepharose Fast Flow; Cytiva, Medemblik, the Netherlands) and eluted using 50 mM Tris (pH 7-8), 300 mM NaCl and 300 mM imidazole. The buffer was exchanged to PBS by overnight dialysis using regenerated cellulose (Spectrum Chemical, Gardena, CA, USA) and proteins were concentrated with Amicon ultrafilters (EMD Millipore, Darmstadt, Germany) and stored at -80°C until use. Protein integrity and purity was assessed using sodium dodecyl sulphate polyacrylamide gel electrophoresis (SDS-PAGE).

### Protein micro-arrays

Protein micro-arrays were performed as described previously.^24,25^ Proteins were mixed with 2× sciSPOT Protein D1 printing buffer (Scienion, Berlin, Germany) and spotted in duplicate in three drops of 333 pL each on 24-pad nitrocellulose-coated slides (ONCYTE AVID, GraceBio Labs, Bend, OR, USA using a sciFLEXARRAYER S12 (Scienion, Berlin, Germany). Slides were dried overnight and stored at room temperature in a low humidity environment. Slides were blocked with Blotto in TBS (ThermoFisher, Breda, The Netherlands) for 1 hour at 37°C then washed 6x using TBS + 0.1% Tween-20. 1:20 diluted sera were used for screening purposes and 3-fold dilution curves starting at 1:20 up to 1:43,740 were used for titre determinations. Sera diluted in Blotto containing 0.1% Surfact-Amps20 (ThermoFisher, Breda, The Netherlands) were added to the slides and incubated 1 hour at 37°C. Slides were washed and incubated with 1:1000 goat anti-human IgG, F(ab’)2 fragment specific, Alexa Fluor 647-conjugated (Jackson Immuno Research, West Grove, PA, USA) diluted in Blotto containing 0.1% Surfact-Amps20 for 1 hour at 37°C. Slides were washed again and then rinsed with MilliQ water and dried. Fluorescent signal was measured using a Tecan Power Scanner (Tecan, Männedorf, Switzerland). Fluorescence intensity was converted to median fluorescent intensity (MFI) values using Scanarray Express version 4.0.0.0004 (PerkinElmer, Rotterdam, the Netherlands). Assay performance and stability was confirmed using the technical duplicates, by use of positive and negative control antigens and via a positive control serum which was titrated in every run.

### Plaque-reduction neutralization tests

Plaque-reduction neutralization tests (PRNTs) were performed as previously described.^24^ A549 cells maintained in Dulbecco’s Modified Eagle Medium (DMEM) supplemented with pyruvate, 10% foetal bovine serum (FBS) and 100 U/mL of penicillin–streptomycin (culture medium) were seeded in 24-well cell culture plates at a density of 5 x 10^4^ cells per well. Sera were diluted 1:8 in A549 infection medium (2% FBS), heat-inactivated for 30 minutes at 56°C and stored overnight at 4°C. The next day, sera were diluted using 2-fold dilutions starting at 1:8 up to 1:2048 in infection medium. Suspension of WNV lineage 2 (strain B956), WNV lineage 1 (strain NY99) or USUV was mixed with the serum dilutions and incubated for 1 hour at 37°C. The resulting inoculum contained 70 PFU/mL and was added on the A549 cells in duplicate and incubated for 1 hour at 37°C, 5% CO_2._ Cells were overlayed using 1% AviCell diluted in infection medium and incubated for 5 days (WNV lineage 2 and USUV) or 4 days (WNV lineage 1) at 37°C, 5% CO_2_. Cells were fixed in 10% paraformaldehyde overnight (WNV) or for 1 hour (USUV) and stained using 1% crystal violet in 20% ethanol. Plaques were counted manually. Endpoint titres were calculated as the highest serum dilution showing a ≥50% reduction in plaque counts compared to wells without serum. Wells without serum typically contained 30-40 plaques. No serum, no virus and cell only controls were included on every plate, results were validated and titres calculated based on within-plate controls.

### Data analysis

For screening purposes, samples were considered positive on the protein micro-array when the average MFI of the duplicate spots was >45,000.^24,26^ For IgG titre determination, half-maximal effective concentration (EC50) values were calculated only if the MFI of the highest serum concentration was >45,000. The lowest detectable EC50 titre is 20.

Probable WNV infections were defined as an EC50 titre on the protein micro-array for any WNV NS1 antigen which was ≥4-fold higher than the EC50 titre for USUV NS1. Probable USUV infections were defined as an EC50 titre for USUV NS1 which was ≥4-fold higher than for any WNV NS1 antigen. All samples with less than a 4-fold EC50 titre difference between USUV NS1 and any of the WNV NS1 antigens were categorized as WNV/USUV. A less strict categorization was applied for the distinction between WNV lineage 1 or 2, to save material during our PRNT testing approach as lineage distinction was not a primary aim of this study. A ≥2-fold EC50 titre difference between the WNV lineage 1 antigen and any of the WNV lineage 2 antigens led to categorization as WNV lineage 1 or 2, while samples with less than a 2-fold titre difference were categorized as WNV/USUV.

For all probable cases, serum samples from earlier rounds (PICO-1 to PICO-10) were selected and used for protein micro-array titration to look for the time (period) of seroconversion and the two subsequent timepoints (PICO-12 and -13) to study antibody kinetics. When seroconversion was noted, samples from earlier rounds were not tested. In this study, the transmission season for WNV and USUV was defined as the months in which there is vector activity (Culex spp.) from May 1st to November 30th.^9^

In alignment with the EU laboratory criteria for WNV case confirmation, probable cases were called confirmed when serological data (micro-array) were confirmed by virus neutralization.^27^ Because this study did not aim to diagnose acute infections, IgM responses were not determined in this study. Sera were tested with a side-by-side confirmatory PRNT setup against USUV and WNV lineage 2 when categorized as probable USUV, probable WNV/USUV or probable WNV lineage 2 infection based on the protein micro-array. Samples were tested against USUV and WNV lineage 1 when categorized as probable WNV lineage 1 infection and tested for all three viruses when there was less than a 2-fold titre difference between both lineages on the protein micro-array. Confirmed WNV infections were defined as samples with a PRNT titre for only WNV (at least 1:8) or ≥4x higher than USUV PRNT titre, irrespective of the protein micro-array results. Confirmed USUV infections had a PRNT titre for only USUV (at least 1:8) or ≥4x higher than WNV PRNT titre, irrespective of the protein micro-array results. Samples were noted as confirmed WNV/USUV when there was a PRNT titre of at least 1:8 for both with a titre difference <4x, irrespective of the protein micro-array results.

The probable and confirmed WNV, USUV and WNV/USUV seroprevalence (and 95% confidence interval (CI)) in the general Dutch population in fall/winter 2023 (PICO-11) was estimated overall and for the probable seroprevalence outcomes also by sex and by age. Additionally, the weighted cumulative infection incidence (and 95% CI) of WNV and USUV up till fall/winter 2023 (PICO-11), as confirmed by seroconversion in a specific time window, was calculated for the general Dutch population as well as by relevant travel history. The national sample (n=4063) in PICO-11 was used as reference population, and the survey design was taken into account (with municipality as a unit of clustering) in all analyses. Post-stratification weights (per study sample) were incorporated using a set of sociodemographic characteristics (age, sex, ethnic background and urbanization degree) to match the Dutch population distribution following census data (of 1 January 2020) from the Statistics Netherlands. Subsequently, estimates were extrapolated to the total number (and 95% CI) of persons in the general Dutch population, using the same census data. The Wald method was used to establish confidence intervals. The aforementioned estimations were repeated in a sensitivity analysis including only participants that were part of the study since the first or second study round. For the probable seroprevalence outcomes by sex and age, potential differences were evaluated using p-values derived from Rao-Scott design-adjusted chi-square tests, and those resulting from design-adjusted F-tests using survey-weighted linear regression, respectively.

Figure 1 contains an overview of the data analysis workflow. All data analysis was performed in Rstudio (version 4.4.0) and SAS (version 9.4., SAS Institute Inc., USA) (for weighting and weighted estimates) and visualization was performed in Rstudio, Graphpad Prism (version 10.5), SankeyMATIC (SankeyMATIC.com), QGIS (version 3.40.5-Bratislava) and Adobe Illustrator (version 28.4.1).

**Figure 1.**
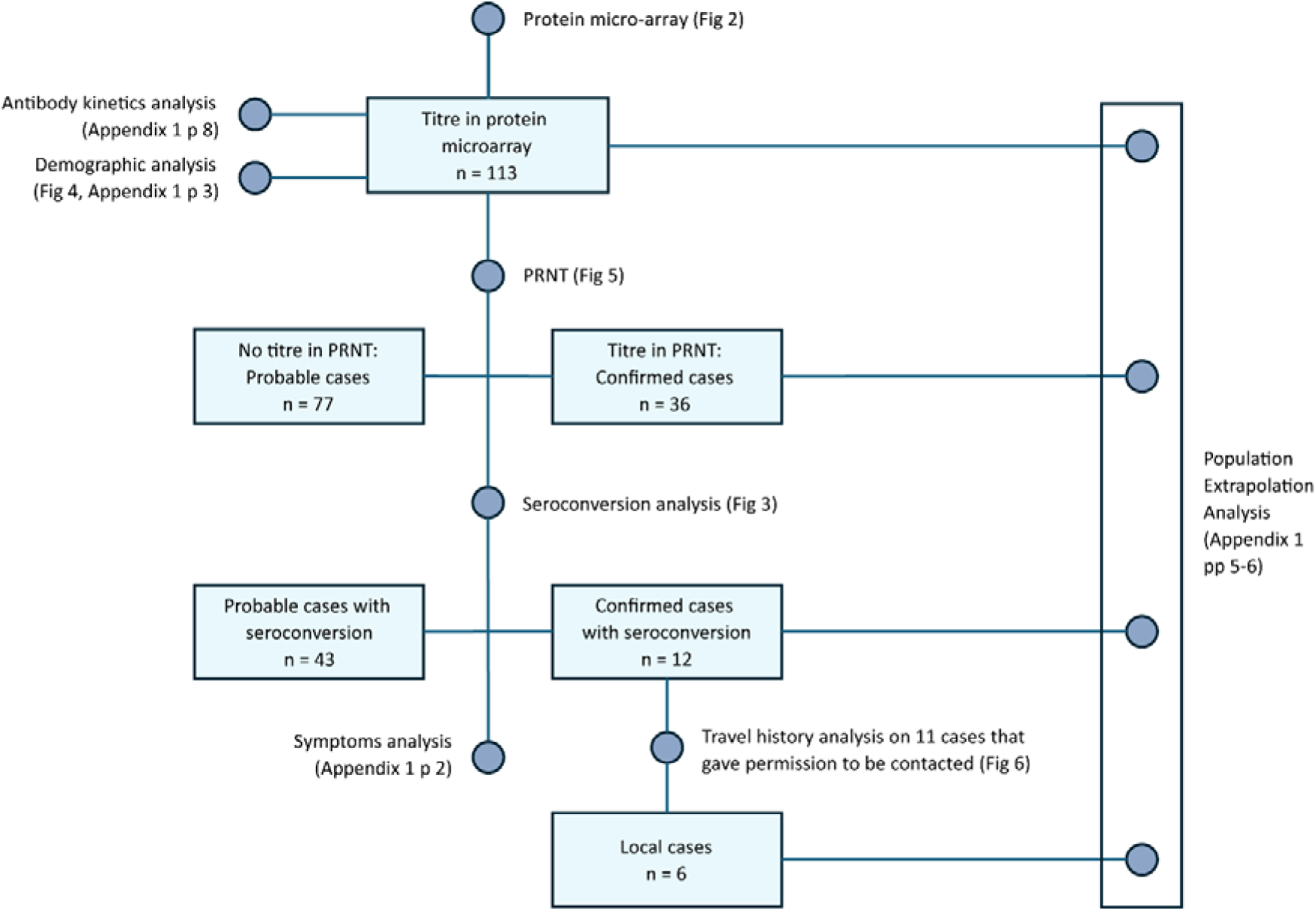
Flowchart showing the data analysis flow and related figures. Abbreviations: n, number; PRNT, plaque reduction neutralization test.

## Results

4977 participants donated a finger stick blood sample between 01-11-2023 and 24-01-2024 (PICO-11) of which 4281 could be included (e.g. due to available serum volume) to screen for the presence of WNV and/or USUV-specific IgG against NS1 by protein micro-array. After a single-point dilution screening, 122 of 4281 (2.8%) sera had an MFI signal above the threshold for at least one of the three antigens. Following titration, 113 of 122 (93%) sera were identified as probable infections and a titre (EC_50_) was determined for the reactive antigens (Figure 1). The 113 titrated sera were categorized in three groups based on observed reactivity patterns: probable WNV infection (n=34, 30%), probable USUV infection (n=54, 48%) and probable WNV/USUV (n=25, 22%) (Figure 2). Twenty-nine of 34 (85%) probable WNV infections had a titre only for WNV lineage 2, two (6%) only for lineage 1, and three (9%) had similar titres (≤2x difference) for WNV lineage 1 and 2 (Appendix 1 p 7).

**Figure 2.**
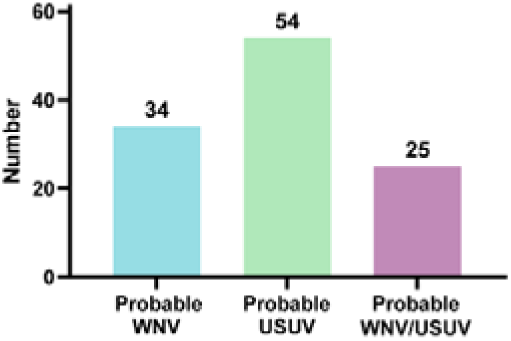
Breakdown of NS1-IgG reactive sera in WNV/USUV micro-array. Y-axis: number of participants positive after two independent measurements: the initial one-point dilution screening and subsequent titration (total n=113). X-axis: Probable WNV case: WNV titre ≥4x higher than USUV titre (n = 34, 85%). Probable USUV case: USUV titre ≥4x higher than WNV titre (n=54, 48%). Probable WNV/USUV case: titre difference less than 4x (n=25, 22%). Abbreviations: WNV, West Nile virus; USUV, Usutu virus; NS1, non-structural protein 1.

To identify possible autochthonous WNV cases, defined as WNV IgG seroconversion with absence of travel to WNV endemic regions, and to enhance our insights into WNV and USUV antibody kinetics, the 113 participants that were positive in the micro-array for WNV and/or USUV NS1 antibodies were individually matched to their corresponding samples collected earlier (PICO-1 to PICO-10) and later (PICO-12 and 13) (Appendix 1 p 7).

To determine the approximate period of infection (period in which seroconversion took place), we tracked these probable cases back in time up to their first negative sample. Fifty-five of 113 probable cases seroconverted for IgG within the study period (WNV n=20, USUV n=28, WNV/USUV n=7). Across all cases, 19 likely seroconverted during the 2023 transmission season, 12 during the 2022 transmission season, three during the 2021 transmission season and seven cases likely seroconverted during the 2020 transmission season (Figure 3). For an additional 14 cases the larger part of the seroconversion window fell outside the mosquito transmission season in the Netherlands,^1^ yet never completely. Ten of the 15 probable cases that participated since PICO-1 (spring 2020), were already positive at the start of the study (WNV n=4, USUV n=5, WNV/USUV n=1).

**Figure 3.**
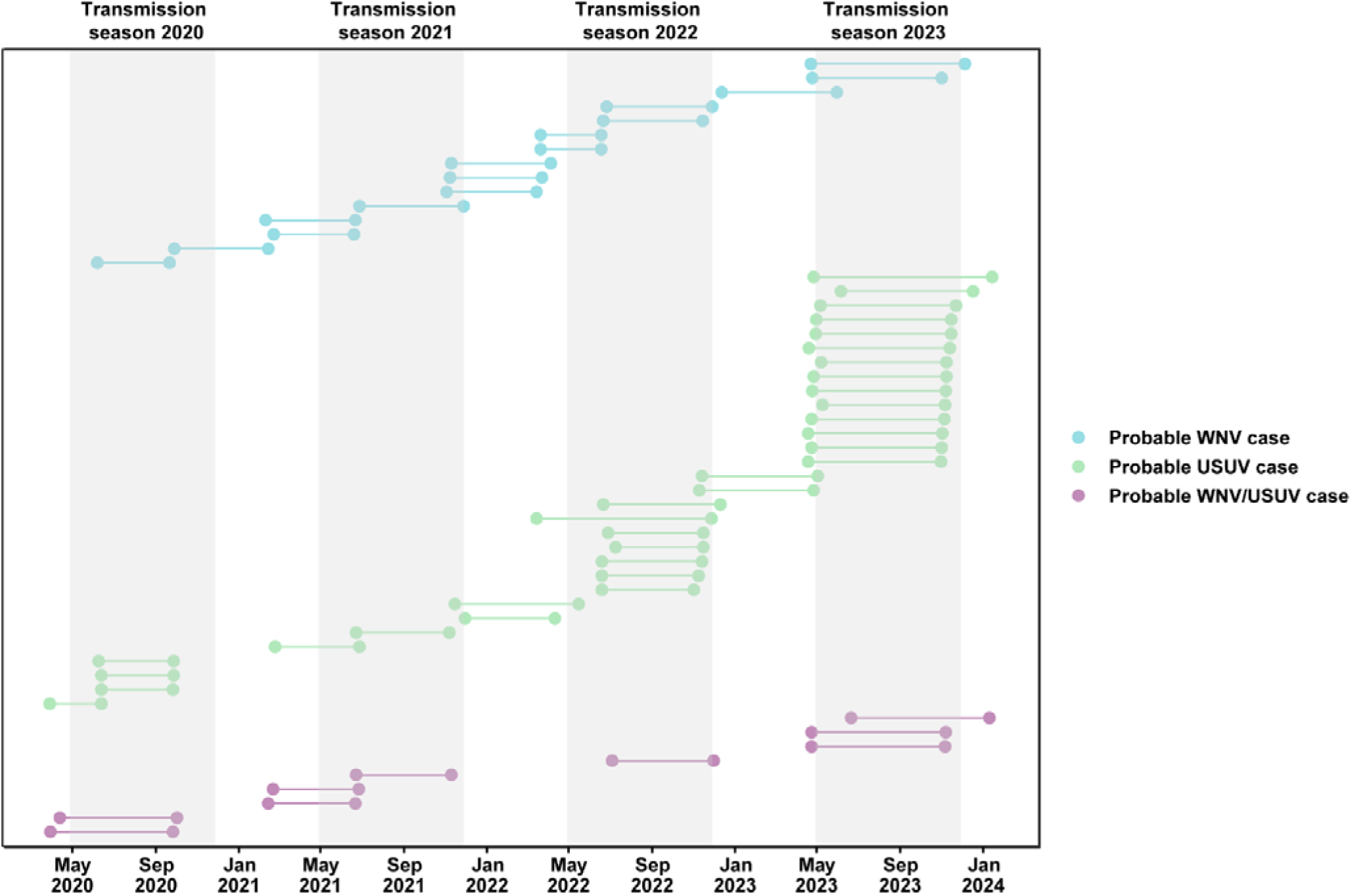
Seroconversion for WNV and USUV in probable cases in a longitudinal cohort (n=55). Seroconversions are shown for probable cases of WNV, USUV and WNV/USUV cases. Connected dots represent the date of the last IgG negative sample (left) and the first IgG positive sample (right) measured for each case. Outcomes are noted as probable WNV/USUV when the EC_50_ titre difference for both viruses is <4x. The light grey areas represent the time windows of the WNV and USUV transmission seasons of 2020, 2021, 2022 and 2023 as defined between May 1^st^ and November 30^th^, when *Culex* mosquitoes are active.^9^ Abbreviations: WNV, West Nile virus; USUV, Usutu virus; EC_50_, half-maximal effective concentration.

To complement the longitudinal retrospective sero-monitoring of positive participants from PICO-11 to PICO-1, we tested all available samples at later timepoints (spring and fall of 2024; PICO-12 and -13) (Appendix 1 p 8). The results demonstrated that IgG antibodies against both WNV and USUV NS1 remained detectable for a long period, up to four years (study end) in some cases.

Questionnaire information was used to gain information on the occurrence of symptoms within the seroconversion period. Although the questionnaires were formulated for COVID-19-related symptoms during the acute phase of the pandemic and respiratory infections in general thereafter, several relevant questions could be included. Twenty out of 55 probable cases reported at least one symptom during the seroconversion period: four out of 15 probable WNV infections, 11 out of 31 probable USUV infections and five out of 9 probable WNV/USUV infections. Three probable cases contacted their general practitioner about their symptoms; none were admitted to the hospital. Notable symptoms possibly in relation to known human clinical manifestation of WNV and USUV infections^28^ included general malaise (n=6), dizziness (n=1), headache (n=8), fever (n=9), muscle ache (n=7) and fatigue (n=3) (Appendix 1 p 2).

There were no significant differences in male/female ratio or age between the three categories based on protein micro-array results versus the negative population, except for a significantly higher age of the probable WNV-infected participants (p<0.004) (Appendix 1 p 3). The place of residence of all 113 probable cases is shown in Figure 4.

**Figure 4.**
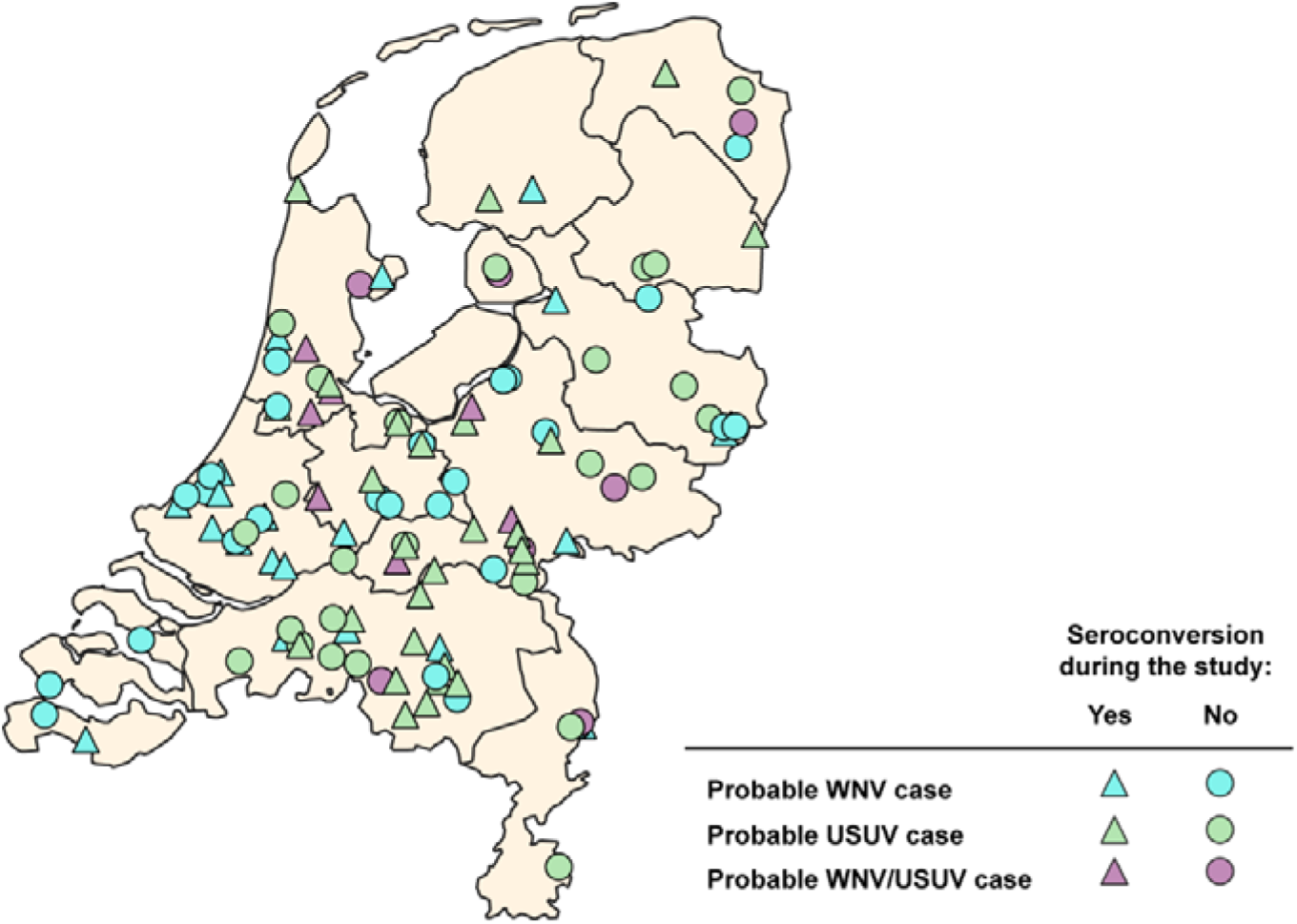
Place of residence for all probable WNV and/or USUV cases, by seroconversion during the study period. The place of residence (in PICO-11) for probable cases is shown on the map of the Netherlands. The place of residence of cases that seroconverted during the study period is shown as triangles. Lines represent the border of provinces. Abbreviations: WNV, West Nile virus; USUV, Usutu virus.

Since orthoflavivirus infection is known to induce an IgG response which can be cross-reactive between members of the orthoflavivirus genus, the gold standard for serology-based confirmation of infection is comparative plaque reduction neutralization testing (PRNT).^27^ To confirm the probable cases, the sera of the 113 probable WNV and/or USUV cases were tested in a comparative PRNT setup against USUV and WNV. Depending on the protein-microarray results, USUV was compared with WNV lineage 2 (n=108), WNV lineage 1 (n=2) or both (n=3). From 34 probable WNV cases, four could be confirmed as WNV cases and two as USUV cases (Figure 5a, Appendix 1 p 8). From 54 probable USUV cases, ten could be confirmed as USUV cases, four as WNV cases and two gave an ambiguous result (WNV/USUV). From the 25 WNV/USUV cases, 10 were confirmed as WNV cases, three as USUV cases and one case remained ambiguous. The remaining 77 samples were negative in the PRNT. The seroconversion period could be identified for six of the 18 confirmed WNV cases, for four of the 15 confirmed USUV cases and for two of the three confirmed WNV/USUV cases (Figure 5b).

**Figure 5.**
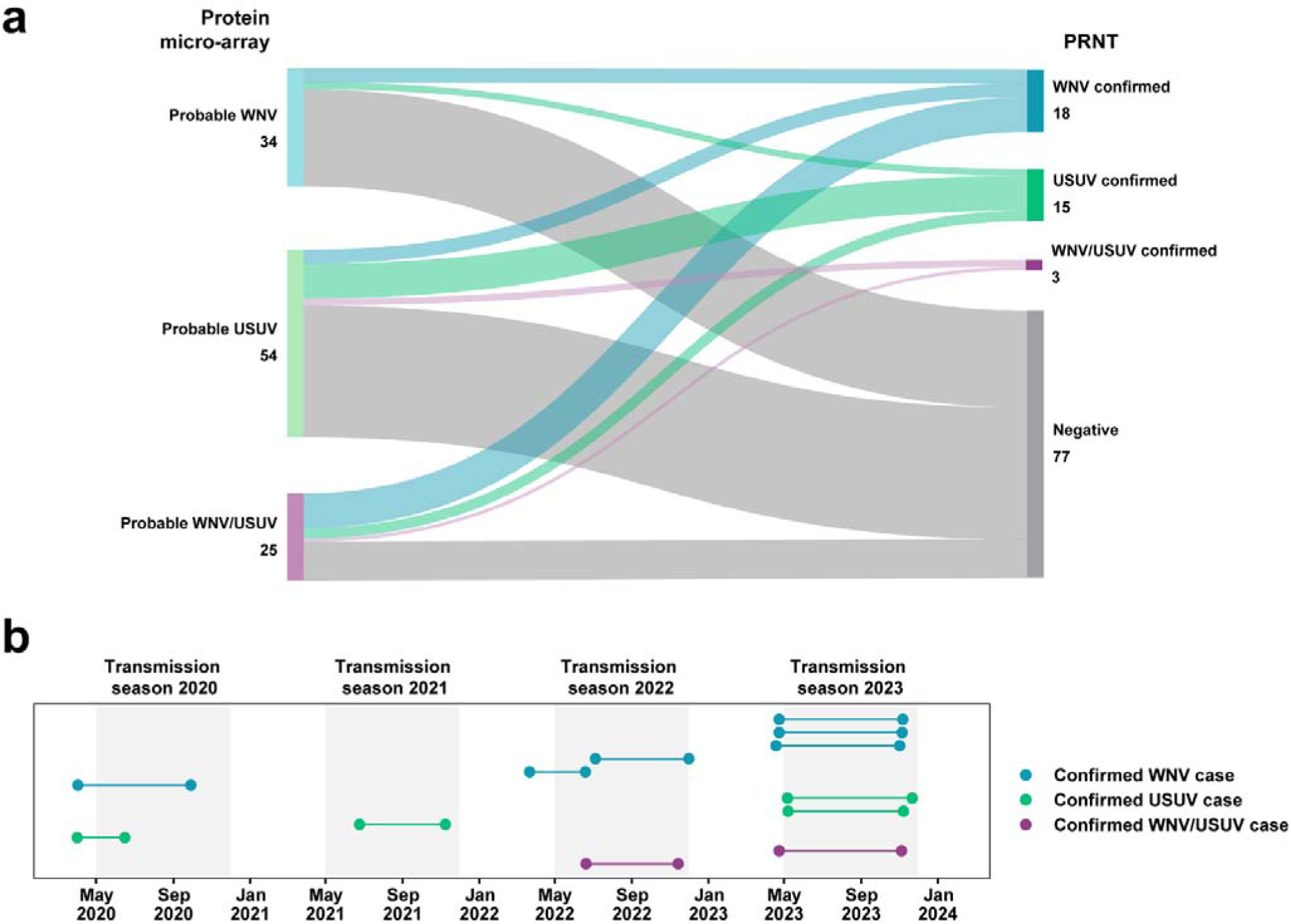
Outcome of confirmatory PRNT assays compared to the protein micro-array results. Outcomes are noted as confirmed WNV/USUV when the PRNT titre difference for both viruses is <4x. (a) Sankey-plot comparing the protein micro-array to the PRNT outcome. (b) Seroconversion in confirmed WNV, USUV and WNV/USUV cases. Connected dots represent the date of the last IgG negative sample (left) and the first IgG positive sample (right) measured for each case. The light grey areas represent the time windows of the WNV and USUV transmission seasons of 2020, 2021, 2022 and 2023 as defined between May 1^st^ and November 30^th^, when *Culex* mosquitoes are active.^9^ Abbreviations: WNV, West Nile virus; USUV, Usutu virus; PRNT, plaque reduction neutralization test.

To determine whether the observed confirmed seroconversions could be the result of autochthonous transmission of WNV and USUV, the confirmed cases were asked for their travel history during the period of seroconversion. Eleven of 12 confirmed seroconverters had given consent to be contacted. Two of four USUV cases (seroconversion in 2021 and 2023), one of six WNV cases (seroconversion 2022) and one WNV/USUV case (seroconversion 2022) reported no travel outside the Netherlands for the period of seroconversion (Appendix 1 p 4). Two WNV cases had travelled to countries with no known WNV circulation at that time (Belgium, and Scotland, United Kingdom). All others had travelled abroad to countries with known WNV and/or USUV circulation. All but one case resided in Dutch provinces with evidence for WNV and/or USUV circulation (Figure 6 and Appendix 1 p 4).

**Figure 6.**
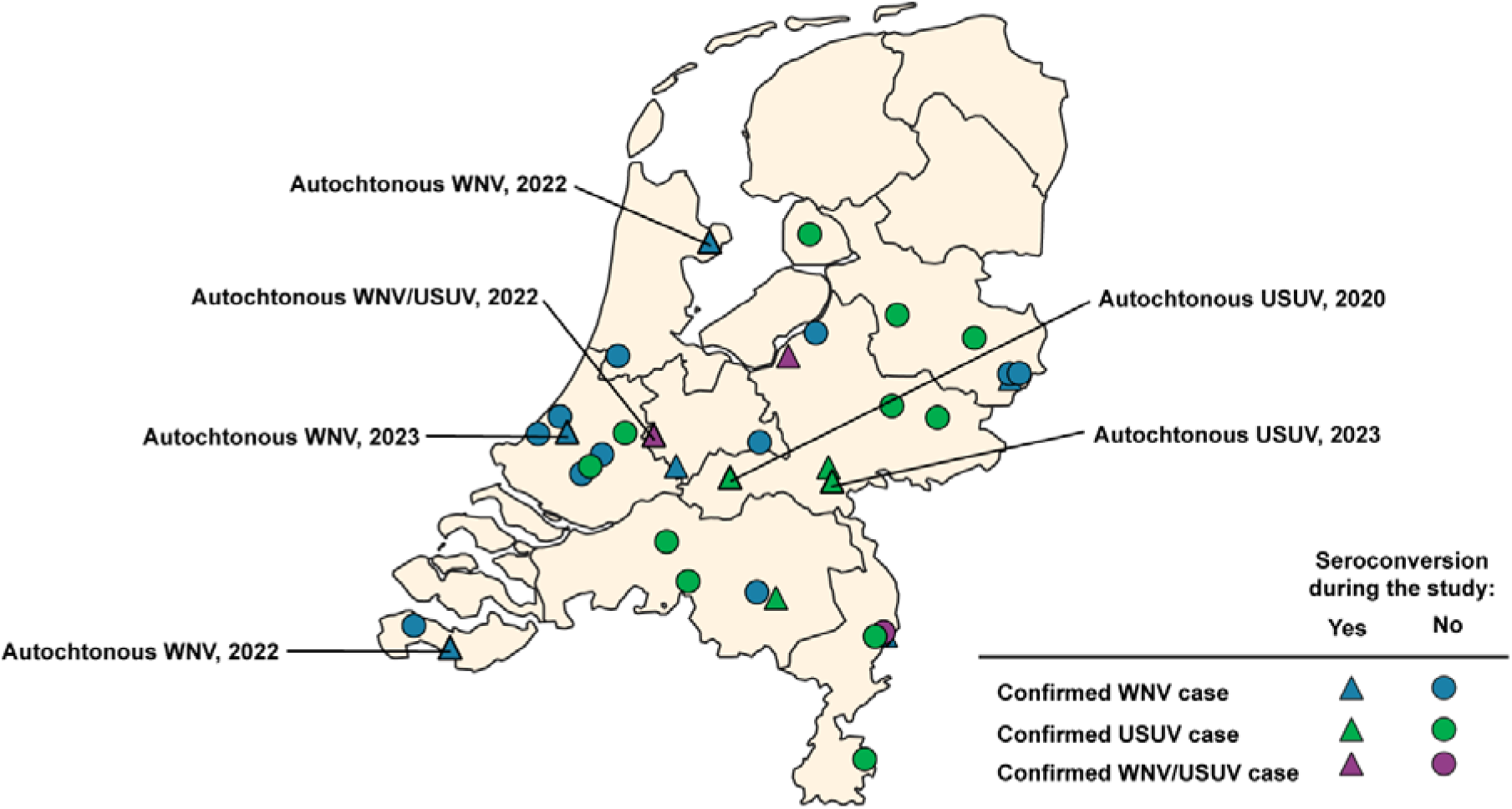
Place of residence for confirmed WNV and/or USUV cases, by seroconversion during the study. The place of residence for PRNT confirmed cases is shown on the map of the Netherlands. The place of residence of cases that seroconverted during the study period is shown as triangles. Lines represent the border of provinces. Autochthonous cases are indicated. Abbreviations: WNV, West Nile virus; USUV, Usutu virus; PRNT, plaque reduction neutralization test.

We estimated nationwide seroprevalence based on various data sets: confirmed cases (PRNT) only, confirmed and probable cases (protein micro-array), with or without ambiguous WNV/USUV cases, and with or without cases with relevant travel history (Appendix 1 p 5). The weighted seroprevalence of probable WNV and USUV cases in the Netherlands in fall/winter 2023 was 0.50% (0.27-0.74%) and 1.33% (0.89-1.78%), respectively. The weighted seroprevalence of confirmed cases only was 0.54% (0.21-0.87%) for WNV and 0.25% (0.09-0.40%) for USUV.

Based on seroconversion during the study period (up till fall/winter 2023), the weighted cumulative incidence in 2020-2023 of WNV and USUV infection in the Dutch population was 0.09% (0.01-0.16%) and 0.06% (0.00-0.13%), respectively. Focussing on locally acquired confirmed infections only during the study period (up till fall/winter 2023), we estimated a weighted cumulative incidence in 2020-2023 of 0.06% (95% CI 0-0.12%) for WNV infection, and of 0.04% (95% CI 0-0.09%) for USUV infection (Appendix 1 p 5). Sensitivity analyses using only participants who were included from the beginning of the study period yielded similar results (Appendix 1 p 6).

## Discussion

Since WNV and USUV were first detected in the Netherlands in 2020 and 2016 respectively, several ecological studies have expanded our knowledge on the presence of these viruses this far north-westward in Europe.^9,10^ The current study describes a unique comprehensive serological screening with longitudinal follow-up for WNV and USUV infection in humans, using a nationwide population-based cohort^20^ and an extensively validated multiplex serological tool in combination with PRNT.^24,25^ The longitudinal approach enabled the identification of (the period of) seroconversion and insight into WNV and USUV IgG longevity while the study design allowed for the identification of autochthonous cases. The study identified 113 probable cases, of which 55 seroconverted during the study, and 36 confirmed cases, among which 12 cases seroconverted. Six of the 12 confirmed cases with seroconversion during the study are most likely locally acquired infections.

Seroprevalence estimates for the overall Dutch population were 0.50% (0.27-0.74%) for WNV and 1.33% (0.89-1.78%) for USUV. Weighted cumulative infection incidence in 2020-2023 during the study period was 0.09% (0.01-0.16%) for WNV and 0.06% (0.00-0.13%) for USUV in the most conservative approach based on PRNT-confirmed cases only. Seroprevalence studies in Europe for comparison are very limited, and positivity rates differ per country, cohort type and time period. In southern France, a 1.2% WNV and 3% USUV seroprevalence was observed in outpatients visiting university hospitals (by microneutralization, 2016-2020).^29^ Studies in blood donors showed a seroprevalence of 0.97% and 0.31% in France (by ELISA and microneutralization respectively, 2021-2022), 0.3% in southern Italy (by PRNT, 2023-2024) and 0.3% in Hamburg, Germany (by microneutralization, 2023).^30–32^

We defined autochthonous infection as IgG seroconversion in absence of travel outside the Netherlands or only to regions without reported WNV or USUV circulation in the period of seroconversion. This was assessed for the 12 confirmed cases and based on recall. Recall inaccuracy might affect the reported history of travel since study participants were contacted up to five years after the period of seroconversion, although most persons were very well aware of this during the study period, i.e., during and shortly after COVID-19 pandemic. For three WNV and four USUV confirmed cases, possible infection during travel could not be excluded. However, for three WNV, two USUV and one WNV/USUV confirmed case, it is highly likely that the infection took place in the Netherlands. Although two of those WNV cases reported travel outside the Netherlands, this was to destinations (Belgium, Scotland) without reported WNV circulation at the time.^6^

Autochthonous human WNV cases in the Netherlands have only been reported in 2020, and very recently, after conclusion of this study, in 2026 with 55 confirmed autochthonous cases. Our study shows that human WNV circulation was ongoing and infections were unrecognized in the Netherlands in the intermittent period 2022-2023, which is in line with previous ecological observations of enzootic circulation in this period.^9,10^ Five of six confirmed WNV cases in our study resided in regions with evidence for WNV circulation based on studies in mosquitoes, birds, horses, rodents and/or wild boars (Appendix 1 p 4). For USUV, the only evidence for human infections in the Netherlands was from a limited molecular study in blood donors and a serological screening of bird ringers.^14,15^ However, ecological surveillance has shown widespread circulation in birds,^10^ including in the regions of residence of the four confirmed human cases (Appendix 1 p 4).

In contrast to other studies presenting seroprevalence estimates in specific cohorts, our study set-up allowed for a weighted extrapolation analysis to the national level. The weighted nationwide cumulative incidence for locally acquired WNV and USUV infection during the study period was estimated at 0.06% (95% CI 0-0.12%) and 0.04% (95% CI 0-0.09%), respectively. The uncertainty of these estimates is high due to the small number of positive cases in the study cohort. However, considering a population size of 17.3 million in 2020 and the percentage of infected cases that develop WNV disease (approximately 20%) or WNV neurological disease (less than 1%),^28^ it appears likely that clinical WNV infections were severely underdiagnosed in the Netherlands even in the group with neurological manifestation that requires hospitalization. The same is likely the case for USUV infections. However, the percentage of USUV infections that require medical care is considered very low in comparison to WNV infections. Hospitalization is mostly observed in cases with underlying medical issues.^33^ A timely and proper diagnosis supports patient management and informs public health authorities to assess the risk and need for control measures.

Demographic analysis outcomes in the current study were in line with what was previously observed for WNV,^34^ namely higher age for WNV cases. While this is the first comprehensive study where demographic analysis could be performed for USUV, those cases did not have significantly different demographics compared to the negative group. While 20 out of 55 probable cases reported symptoms during the period of seroconversion, it is likely that part of those are related to other conditions or infections, indicating that our positive population is largely asymptomatic.

We have previously shown that the protein micro-array is reliable, sensitive and specific for detection of symptomatic patients with a recent infection (< 1 month) and that cohort-dependent differences in outcomes between the micro-array and PRNT exist.^8,24,25^ Here, we studied a general population cohort with a potential mixture of recent and less recent, and mostly mild or asymptomatic infections, and observed that only part of the probable cases could be confirmed. A possible explanation could be the waning of (neutralizing) antibody levels. Moreover, fundamental differences exist between the two assays. For example, epitope specificity can significantly impact whether antibodies are also able to mediate virus neutralization.^35^ Consequently, protein microarray results provide additional (complementary) and valuable information on possible WNV and USUV exposures.

Interestingly, confirmed WNV cases more frequently had an ambiguous outcome on the protein micro-array than the confirmed USUV cases. This could indicate a difference in assay performance, possibly related to differences in the ability of WNV infection to induce antibodies against USUV and vice versa. Such differences have been described by others.^36^ Due to volume constraints, most sera were tested only for WNV lineage 2 in the PRNT. While this is the lineage known to circulate in the Netherlands in 2020, there is a small chance that we missed confirming WNV lineage 1 infections by PRNT, which could have been acquired during travel or from unrecognized circulation in the Netherlands.

Information on the dynamics and longevity of IgG antibody responses against WNV is scarce while longevity studies for USUV have not been described. Our data shows that human IgG responses against USUV can persist up to four years (total study duration), with dynamics similar to WNV. A few studies have assessed the dynamics of WNV-specific antibodies after infection but in specific cohorts. In mostly asymptomatic blood donors, IgG antibodies to WNV were shown to persist typically for up to a year after acute infection,^37^ while one study showed IgG persistence up to five years.^38^ The observed persistence of USUV IgG for at least four years in our study is in line with what is known for WNV and other orthoflaviruses.^39^ While cross-protection between WNV and USUV has been demonstrated in birds and animal models,^40,41^ corresponding data on humans is absent. The USUV IgG seroprevalence determined in this study could provide essential information for studies investigating potential cross-protection of USUV infection to severe clinical manifestation of WNV. Our data will serve as baseline data for future serological surveillance in a context where WNV and USUV will continue to emerge. Knowledge on the longevity of IgG responses is highly valuable for a proper interpretation of such studies.

This study has several limitations. While our study design entails a representative selection of the general Dutch population, some selective response was notable, e.g. participation of relatively more native Dutch, middle-aged inhabitants, women and those residing in areas with a lower level of urbanization. Yet, the weighted analysis allowed for adjustment of important factors which deviated from the distribution on the national level. Not all participants were included since the start of the study, therefore we may have missed seroconversions for participants that enrolled later. However, a sensitivity analysis was performed showing that this has likely had minimal effect on our estimates. Participants reported symptoms during each study round, however since they were focused on respiratory infections, symptoms that are relevant to this study may be missing.

Using a unique longitudinal serological nationwide cohort study, we studied the incidence of local WNV infection in the general population of the Netherlands during a four-year period with evidence for WNV emergence. We identified autochthonous WNV and USUV infections, determined the overall seroprevalence for both viruses and quantified the weighted cumulative incidence based on local infections during this four-year period. Our study shows that infections with newly emerging pathogens like WNV went undetected and suggests that clinical cases were most likely vastly underdiagnosed. It highlights the need for an increased awareness and vigilance among physicians to identify clinical cases and the need for continued paired syndromic-laboratory surveillance for WNV and USUV in the Netherlands to ensure proper patient and public health management. Our study underscores the high value of the versatile use of long-term, longitudinal and population-based cohorts to address emerging public health questions.

## Author contributions

Conceptualization and study design: Chantal Reusken, Johan Reimerink, Marloes Grobben, Gerco den Hartog, Coralie Valle. Methodology: Coralie Valle, Gert-Jan Godeke, Gerco den Hartog, Marloes Grobben. Investigation: Corine Nellestijn, Hanae Abba, Sandhya Shrestha, Gert-Jan Godeke, Bettie Voordouw, Fiona van der Klis, Gerco den Hartog, Eelco Franz, Eric Vos, Dirk Eggink. Data access, verification, management and analysis: Marloes Grobben, Eric Vos. Manuscript draft: Marloes Grobben, Chantal Reusken, Johan Reimerink. Responsible for the decision to submit the manuscript: Chantal Reusken, Johan Reimerink, Gerco den Hartog. Critical revision of manuscript: All authors. Approved the final manuscript: All authors.

## Data Availability

The data presented in this study are available on request from the corresponding author.

## Acknowledgements

We would like to thank all participants of the PICO study. We thank Trung Bui, Patrick Boers and Kelly Breedijk for laboratory support and Gaby Smits, Arginell Girigorie and Kira Thijse for their assistance with the serum samples. The PICO cohort and this study were funded by the ministry of Health, Welfare and Sports (VWS), the Netherlands.

