## Supplementary materials for "Human infection incidence for West Nile virus and Usutu virus in the Netherlands: evidence from a longitudinal nationwide cohort, 2020-2023"

**Table of contents:**

**Supplementary table 1.** Baseline characteristics. . Page 2

**Supplementary table 2.** Symptoms reported during the seroconversion period. Page 2

**Supplementary table 3.** Gender distribution. Page 3

**Supplementary table 4.** Age distribution. Page 3

**Supplementary table 5.** Travel history of WNV and/or USUV cases. Page 4

**Supplementary table 6.** Seroprevalence and weighed infection incidence. Page 5

**Supplementary table 7.** Sensitivity analysis for seroprevalence and weighed infection incidence. Page 6

**Supplementary Figure 1.** Study set-up. Page 7

**Supplementary Figure 2.** Breakdown of probable WNV cases at lineage level. Page 7

**Supplementary Figure 3.** NS1 micro-array IgG titres over time. Page 8

**Supplementary Figure 4.** PRNT titres. Page 8

**References.** Page 9

**Supplementary table 1. Baseline characteristics of all participants included in this study.** The total number of participants included in this study is 4281. Abbreviations: IQR, interquartile range.

| | **All participants included in this study** |
|---|---|
| **Age in years, median (IQR)** | 59 (41-72) |
| **Gender, number (percent)** | |
| Male | 1871 (44%) |
| Female | 2416 (56%) |
| **Region, number (percent)** | |
| Drenthe | 137 (3.2%) |
| Flevoland | 76 (1.78%) |
| Friesland | 217 (5.07%) |
| Gelderland | 667 (15.58%) |
| Groningen | 201 (4.7%) |
| Limburg | 225 (5.26%) |
| Noord-Brabant | 592 (13.83%) |
| Noord-Holland | 674 (15.74%) |
| Overijssel | 341 (7.97%) |
| Utrecht | 325 (7.59%) |
| Zeeland | 159 (3.71%) |
| Zuid-Holland | 667 (15.58%) |

**Supplementary table 2. Symptoms reported by 51 of 55 probable WNV and USUV cases during the seroconversion period.** Results are shown for 51 probable cases (based on protein micro-array) where we observed seroconversion during the study period. For four cases no information regarding symptoms was available. Numbers represent the number of probable cases that reported each symptom. Probable cases were broken down to pathogen level; probable WNV infection (n = 14), probable USUV infection (n = 28) and probable WNV/USUV infection (n = 9). Abbreviations: WNV, West Nile virus; USUV, Usutu virus; n, number.

| | **Probable WNV cases (n)** | **Probable USUV cases (n)** | **Probable WNV/USUV cases (n)** |
|---|---|---|---|
| General malaise | 0 | 4 | 2 |
| Stomach-ache | 0 | 1 | 0 |
| Diarrhoea | 0 | 0 | 0 |
| Dizziness | 0 | 1 | 0 |
| Loss of smell and/or taste | 0 | 1 | 1 |
| Joint pain | 0 | 1 | 1 |
| Cough | 1 | 5 | 2 |
| Headache | 0 | 6 | 2 |
| Sore throat | 1 | 7 | 0 |
| Fever | 2 | 4 | 3 |
| Shortness of breath | 0 | 2 | 1 |
| Runny nose | 2 | 5 | 2 |
| Nausea | 0 | 0 | 0 |
| Painful breathing | 0 | 2 | 0 |
| Pain behind eyes | 0 | 1 | 1 |
| Irritability/confusion | 0 | 1 | 0 |
| Muscle ache | 2 | 4 | 1 |
| Fatigue | 0 | 3 | 0 |

**Supplementary table 3. Weighted proportion of probable cases in the Dutch population by sex.** The weighted estimates of probable WNV includingWNV/USUV cases and of probable USUV including WNV/USUV cases were each compared with the rest of the study population. Differences were evaluated using p-values derived from Roa-Scott design-adjusted chi-square tests. Abbreviations: WNV, West Nile virus; USUV, Usutu virus; CI, confidence interval (Wald).

| Result | Sex | Number of positives/total | Weighted estimate, percent (95% CI) | P-value |
|---|---|---|---|---|
| Probable WNV including WNV/USUV | Male | 31/1777 | 1.08% (0.58-1.59%) | 0.86 |
| | Female | 25/2286 | 1.15% (0.61-1.69%) | |
| Probable USUV including WNV/USUV | Male | 34/1777 | 1.91% (1.04-2.77%) | 0.89 |
| | Female | 44/2286 | 1.99% (1.25-2.72%) | |

**Supplementary table 4. Weighted mean age of probable cases**. The mean age of probable WNV includingWNV/USUV cases and of probable USUV including WNV/USUV cases were each compared with the rest of the study population. Differences were evaluated using p-values derived from design-adjusted F-tests using survey-weighted linear regression. Abbreviations: WNV, West Nile virus; USUV, Usutu virus; CI, confidence interval (Wald).

| **WNV** | | | |
|---|---|---|---|
| **Result** | **Number of participants** | **Age, mean (95% CI)** | **P-value** |
| Probable WNV including WNV/USUV | 56 | 55.8% (50.4-61.3%) | <0.0001 |
| Other participants | 4007 | 41.9% (40.8-43.1%) | |
| **USUV** | | | |
| **Result** | **Number of participants** | **Mean** | **P-value** |
| Probable USUV including WNV/USUV | 78 | 46.1% (41.4-50.7%) | 0.091 |
| Other participants | 3985 | 42% (40.9-43.2%) | |

**Supplementary table 5. Travel history of WNV and/or USUV cases.** Travel history was collected only for confirmed WNV, USUV, or WNV/USUV cases with seroconversion detected during the study period to identify possible autochthonous infections n=11). Region definitions are based on the COROP regions, a standardized geocode system used for data analysis, equivalent to the NUTS-3 regions of the Netherlands. Abbreviations: WNV, West Nile virus; USUV, Usutu virus; PRNT, plaque reduction neutralization test; Nabs, neutralizing antibodies.

| | | | | |
|---|---|---|---|---|
| **USUV cases** | | | | |
| **#** | **Sero-conversion year** | **Travel history in the seroconversion period** | **Place of residence (region, province)** | **Other evidence of circulation in that province (type of evidence, year(s))** |
| 1 | 2020 | No international travel | Zuidwest-Gelderland, Gelderland | Birds (RNA, 2016-2022),[1,2] carnivores (Nabs, 2020-2023),[3] mosquitoes (RNA, 2019-2022,[2] |
| 2 | 2021 | Visited the south of France | Arnhem/Nijmegen, Gelderland | |
| 3 | 2023 | No international travel | Arnhem/Nijmegen, Gelderland | |
| 4 | 2023 | Visited the south of France | Zuidoost-Noord-Brabant, Noord-Brabant | Birds (RNA, 2016-2022),[1,2] mosquitoes (RNA, 2019-2022)[2] |
| **WNV cases** | | | | |
| **#** | **Sero-conversion year** | **Travel history in the seroconversion period** | **Place of residence (region, province)** | **Other evidence of circulation in that province** |
| 1 | 2020 | Visited the Czech Republic | Twente, Overijssel | Human cases (RNA, 2026),[4] Horse* (RNA, 2023),[5] birds (Nabs, 2020, 2022)[2] |
| 2 | 2022 | No international travel | Kop van Noord-Holland, Noord-Holland | Human cases (RNA, 2026),[4] Birds (Nabs 2020, 2022),[2] rodents (Nabs, 2023)[6] |
| 3 | 2022 | Visited Belgium | Zeeuws-Vlaanderen, Zeeland | No data available |
| 4 | 2023 | Visited Scotland | s-Gravenhage, Zuid-Holland | Human cases (RNA, 2026),[4] Horses (Nabs, 2021-2022, 2025),[7,8] carnivore (Nabs, 2020-2023),[3] mosquitoes (RNA, 2025)[8] |
| 5 | 2023 | Visited India and Germany | Noord-Limburg, Limburg | Wild boars (Nabs, 2018-2021)[9] |
| 6 | 2023 | Visited Germany | Utrecht, Utrecht | Human cases (RNA/Nabs, 2020; RNA, 2026),[4,10] birds (RNA, 2020; Nabs, 2020, 2022),[2,11] mosquitoes (RNA, 2020),[11] chickens (WNV Nabs, 2021-2022),[12] rodents (Nabs, 2021)[6] |
| **WNV/USUV cases** | | | | |
| **#** | **Sero-conversion year** | **Travel history in the seroconversion period** | **Place of residence (region, province)** | **Other evidence of circulation in that province** |
| 1 | 2022 | No international travel | Utrecht, Utrecht | Human cases (WNV RNA/Nabs, 2020; WNV RNA, 2026),[4,10] birds (WNV RNA, 2020; WNV Nabs, 2020, 2022; USUV RNA/Nab, 2016-2022),[1,2,11] mosquitoes (WNV RNA, 2020; USUV RNA, 2019-2022),[2,11] chickens (WNV Nabs, 2021-2022),[12] rodents (WNV Nabs, 2021)[6] |
| 2 | 2022 | No information available | Veluwe, Gelderland | Human cases (WNV RNA, 2020; WNV RNA, 2026),[4,10] birds (WNV Nabs, 2020, 2022; USUV RNA, 2016-2022),[1,2] mosquitoes (USUV RNA, 2019-2022),[2] chickens (WNV Nabs, 2021-2022)[12], carnivores (USUV Nabs, 2020-2023)[3] |

**Supplementary table 6. Weighted seroprevalence and infection incidence for WNV and/or USUV, extrapolated to the overall Dutch population.** Countries that were not considered relevant travel history for WNV infection: Belgium and Scotland (United Kingdom). Abbreviations: WNV, West Nile virus; USUV, Usutu virus; NA, not applicable; CI, confidence interval (Wald).

| **WNV seroprevalence** | | | | |
|---|---|---|---|---|
| **Case definition** | **Result** | **Number of positives/total** | **Weighted estimate, percent (95% CI)** | **Estimated number of persons (95% CI)** |
| Probable | WNV + WNV/USUV | 56/4063 | 1.12% (0.76-1.47%) | 193000 (131936-254064) |
| Probable | WNV | 31/4063 | 0.50% (0.27-0.74%) | 87023 (46526-127520) |
| Confirmed | WNV + WNV/USUV | 21/4063 | 0.59% (0.25-0.93%) | 102265 (43648-160882) |
| Confirmed | WNV | 18/4063 | 0.54% (0.21-0.87%) | 92967 (35912-150022) |
| **USUV seroprevalence** | | | | |
| **Case definition** | **Result** | **Number of positives/total** | **Weighted estimate, percent (95% CI)** | **Estimated number of persons (95% CI)** |
| Probable | USUV + WNV/USUV | 78/4063 | 1.95% (1.39-2.50%) | 336859 (241135-432583) |
| Probable | USUV | 53/4063 | 1.33% (0.89-1.78%) | 230881 (154205-307558) |
| Confirmed | USUV + WNV/USUV | 18/4063 | 0.30% (0.14-0.47%) | 52079 (23677-80480) |
| Confirmed | USUV | 15/4063 | 0.25% (0.09-0.40%) | 42780 (16353-69208) |

| **WNV cumulative infection incidence in 2020-2023** | | | | |
|---|---|---|---|---|
| **Result** | **Selection** | **Number of positives/total** | **Weighted estimate, percent (95% CI)** | **Estimated number of persons (95% CI)** |
| WNV + WNV/USUV | all | 8/4063 | 0.12% (0.03-0.20%) | 20206 (5119-35293) |
| WNV | all | 6/4063 | 0.09% (0.01-0.16%) | 14761 (1927-27596) |
| WNV + WNV/USUV | without relevant travel history | 4/4063 | 0.07% (0.00-0.14%) | 11620 (0-23800) |
| WNV | without relevant travel history | 3/4063 | 0.06% (0.00-0.12%) | 9521 (0-20944) |
| **USUV cumulative infection incidence in 2020-2023** | | | | |
| **Result** | **Selection** | **Number of positives/total** | **Weighted estimate, percent (95% CI)** | **Estimated number of persons (95% CI)** |
| USUV + WNV/USUV | all | 6/4063 | 0.10% (0.01-0.18%) | 16479 (2550-30408) |
| USUV | all | 4/4063 | 0.06% (0.00-0.13%) | 11035 (0-22544) |
| USUV + WNV/USUV | without relevant travel history | 3/4063 | 0.05% (0.00-0.11%) | 8288 (0-18482) |
| USUV | without relevant travel history | 2/4063 | 0.04% (0.00-0.09%) | 6190 (0-15495) |

**Supplementary table 7. Sensitivity analysis for weighted seroprevalence and infection incidence for WNV and/or USUV including only participants that were included in study round 1 or 2.** Countries that were not considered relevant travel history for WNV infection: Belgium and Scotland (United Kingdom). Abbreviations: WNV, West Nile virus; USUV, Usutu virus; NA, not applicable; CI, confidence interval (Wald).

| **WNV seroprevalence** | | | | |
|---|---|---|---|---|
| **Case definition** | **Result** | **Number of positives/total** | **Weighted estimate, percent (95% CI)** | **Estimated number of persons (95% CI)** |
| Probable | WNV + WNV/USUV | 36/2786 | 1.15% (0.76-1.54%) | 199166 (131779-266554) |
| Probable | WNV | 17/2786 | 0.42% (0.21-0.63%) | 72022 (35830-108215) |
| Confirmed | WNV + WNV/USUV | 13/2786 | 0.48% (0.15-0.82%) | 83520 (25448-141592) |
| Confirmed | WNV | 13/2786 | 0.48% (0.15-0.82%) | 83520 (25448-141592) |
| **USUV seroprevalence** | | | | |
| **Case definition** | **Result** | **Number of positives/total** | **Weighted estimate, percent (95% CI)** | **Estimated number of persons (95% CI)** |
| Probable | USUV + WNV/USUV | 52/2786 | 2% (1.22-2.78%) | 346690 (211980-481399) |
| Probable | USUV | 33/2786 | 1.27% (0.71-1.83%) | 219546 (122704-316388) |
| Confirmed | USUV + WNV/USUV | 9/2786 | 0.33% (0-0.67%) | 56759 (0-116466) |
| Confirmed | USUV | 9/2786 | 0.33% (0-0.67%) | 56759 (0-116466) |

| **WNV cumulative infection incidence in 2020-2023** | | | | |
|---|---|---|---|---|
| **Result** | **Selection** | **Number of positives/total** | **Weighted estimate, percent (95% CI)** | **Estimated number of persons (95% CI)** |
| WNV + WNV/USUV | all | 6/2786 | 0.11% (0.01-0.21%) | 19182 (1272-37092) |
| WNV | all | 6/2786 | 0.11% (0.01-0.21%) | 19182 (1272-37092) |
| WNV + WNV/USUV | without relevant travel history | 3/2786 | 0.07% (0-0.17%) | 12728 (0-29159) |
| WNV | without relevant travel history | 3/2786 | 0.07% (0-0.17%) | 12728 (0-29159) |
| **USUV cumulative infection incidence in 2020-2023** | | | | |
| **Result** | **Selection** | **Number of positives/total** | **Weighted estimate, percent (95% CI)** | **Estimated number of persons (95% CI)** |
| USUV + WNV/USUV | all | 4/2786 | 0.1% (0-0.2%) | 16801 (0-34028) |
| USUV | all | 4/2786 | 0.1% (0-0.2%) | 16801 (0-34028) |
| USUV + WNV/USUV | without relevant travel history | 2/2786 | 0.05% (0-0.13%) | 9027 (0-22580) |
| USUV | without relevant travel history | 2/2786 | 0.05% (0-0.13%) | 9027 (0-22580) |

a

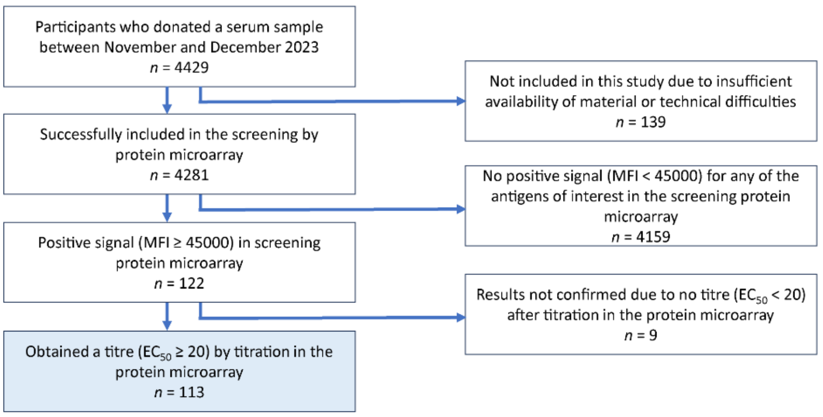


b

| | Sample tested (n) | Earliest sample date this round | Latest sample date this round | Serocon-verted in or after round 10 (n) | Serocon-verted in or after round 7 (n) | Serocon-verted in or after round 4 (n) | Sample not available (n) |
|---|---|---|---|---|---|---|---|
| **PICO 1** | 15 | 1-4-2020 | 22-4-2020 | 19 | 15 | 13 | 51 |
| **PICO 2** | 32 | 10-6-2020 | 30-6-2020 | 19 | 15 | 13 | 34 |
| **PICO 3** | 33 | 24-9-2020 | 20-11-2020 | 19 | 15 | 13 | 33 |
| **PICO 4** | 46 | 12-2-2021 | 26-2-2021 | 19 | 15 | NA | 33 |
| **PICO 5** | 47 | 18-6-2021 | 30-7-2021 | 19 | 15 | NA | 32 |
| **PICO 6** | 78 | 3-11-2021 | 3-12-2021 | 19 | 15 | NA | 1 |
| **PICO 7** | 89 | 16-3-2022 | 18-5-2022 | 19 | NA | NA | 5 |
| **PICO 8** | 87 | 16-6-2022 | 10-8-2022 | 19 | NA | NA | 7 |
| **PICO 9** | 86 | 2-11-2022 | 9-1-2023 | 19 | NA | NA | 8 |
| **PICO 10** | 105 | 19-4-2023 | 30-6-2023 | NA | NA | NA | 8 |
| **PICO 11** | 113 | 1-11-2023 | 24-1-2024 | NA | NA | NA | NA |
| **PICO 12** | 100 | 11-4-2024 | 12-6-2024 | NA | NA | NA | 13 |
| **PICO 13** | 94 | 9-10-2024 | 16-12-2024 | NA | NA | NA | 19 |

c

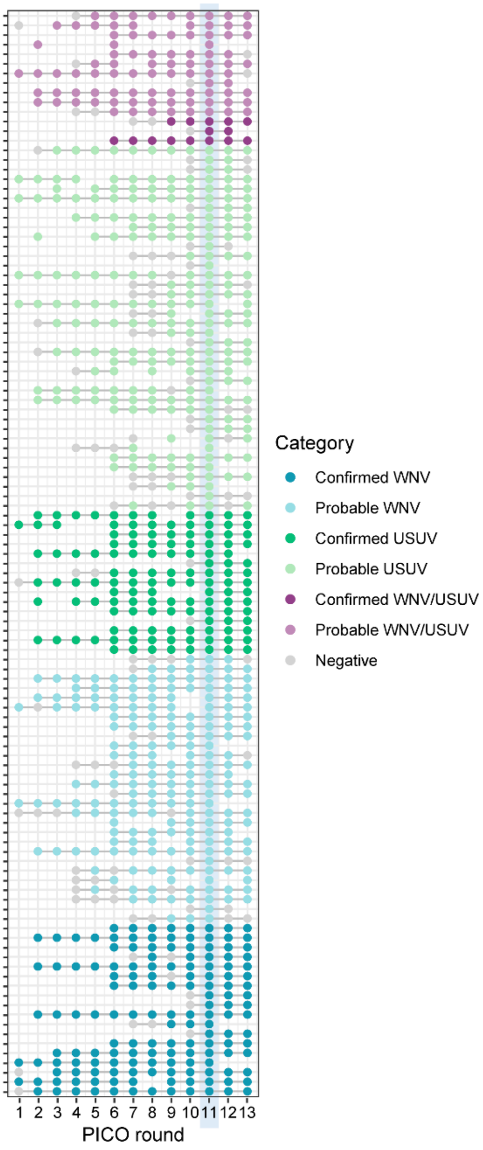


**Supplementary Figure 1. Study set-up.** (a) Flowchart representing the process of testing all available samples from Fall/Winter 2023 (PICO-11, n = 4429) up to the 113 samples identified as positive for WNV and/or USUV by protein micro-array. (b) Timeline of study rounds showing sample availability. New participants were included in rounds 2 and 6. Where seroconversion was noted, samples from earlier rounds were not tested since they are assumed to be negative as well. (c) Overview of sample availability for the 113 positive samples over the different rounds. Colour coding was applied based on the PRNT outcome and if the PRNT was negative, based on the categorization of protein micro-array results in PICO-11. Negative samples are coloured in grey. Abbreviations: PICO: Pienter Corona study; WNV, West Nile virus; USUV, Usutu virus; n, number; $EC_{50}$, half-maximal effective concentration, MFI: median fluorescence intensity; PRNT, plaque reduction neutralization test.

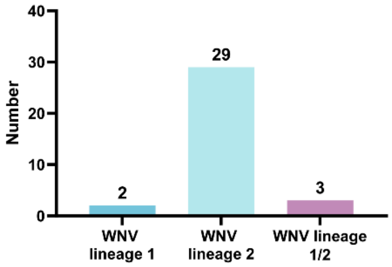


**Supplementary Figure 2. Breakdown of probable WNV cases at lineage level.** Y-axis: number of probable WNV cases positive after two independent measurements: the initial one-point dilution screening and subsequent titration (total n = 34). X-axis: WNV probable cases were categorized as WNV lineage 1 (n=2, 6%) or 2 (n=29, 85%) when there is at least a ≥2x higher titre for one of these lineages. When the titre difference was smaller, the cases were categorized as WNV lineage 1/2 (n=3, 9%). Abbreviations: WNV, West Nile virus.

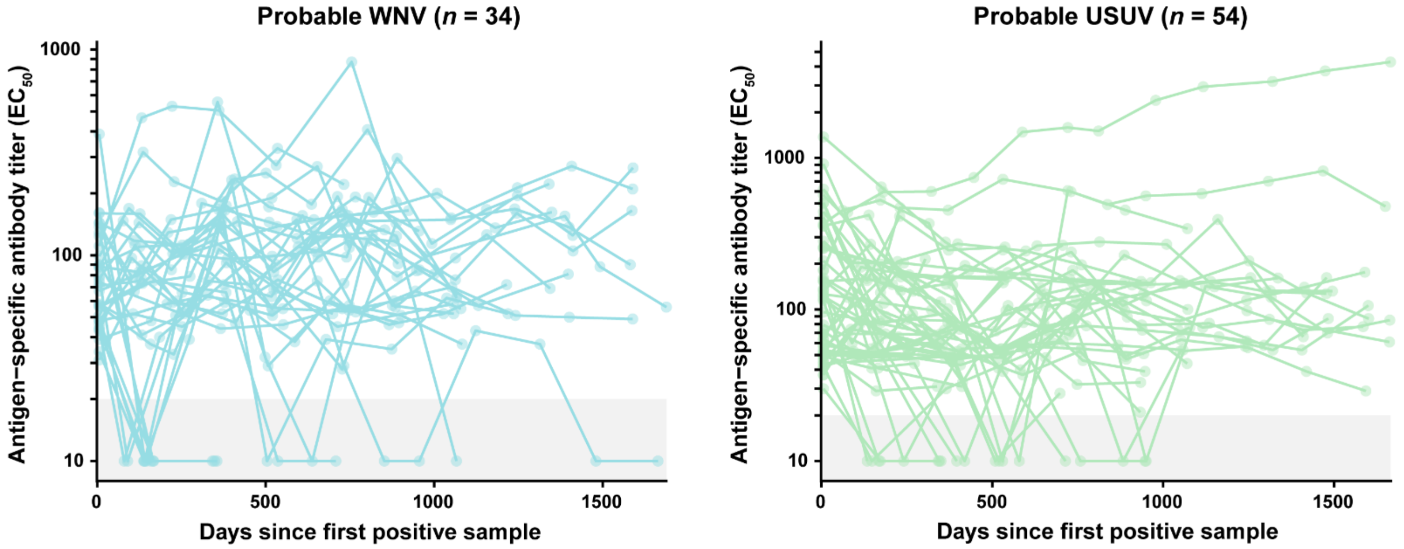


**Supplementary Figure 3. NS1 micro-array IgG titres over time for participants with a probable infection with WNV (n=34) or USUV (n=54).** Y-axis: $EC_{50}$ titre for anti-NS1 IgG. X-axis: days since the first positive sample. All data points for all participants have been plotted, and lines connect the different points belonging to the same individual. Negative datapoints before seroconversion have been excluded to show only data from the first positive sample onwards. Abbreviations: WNV, West Nile virus; USUV, Usutu virus; $EC_{50}$, half-maximal effective concentration.

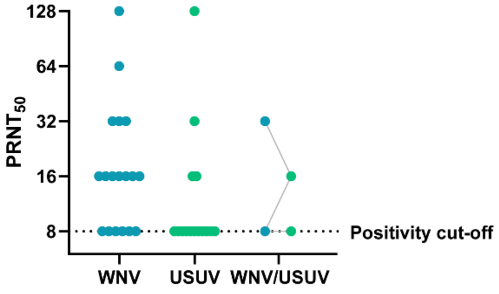


**Supplementary Figure 4. PRNT titres of participants with a confirmed WNV, USUV and WNV/USUV infection.** Y-axis: $PRNT_{50}$ titre. X-axis: categorization of participants as confirmed WNV, USUV and WNV/USUV infection. Titres for WNV are shown in blue and titres for USUV are shown in green. In the WNV/USUV category, grey lines connect the WNV and USUV titre of the same sample. Abbreviations: WNV, West Nile virus; USUV, Usutu virus; PRNT, plaque reduction neutralization test; $PRNT_{50}$, half-maximal plaque reduction titre.
